# Evaluating Predictive Value of *Schistosoma mansoni* Prevalence and Infection Intensity in School-Age Children in Estimating Community-Wide Prevalence in Siaya County, Western Kenya

**DOI:** 10.1101/2024.02.19.24302832

**Authors:** Dollycate Wanja, Maurice R. Odiere, Emmy A. Kavere, Geoffrey Muchiri, Shehu Shagari Awandu

**Affiliations:** Department of Biomedical Sciences, Jaramogi Oginga Odinga University of Science and Technology, Bondo, Kenya; Centre for Global Health and Research, Kenya Medical Research Institute, /, Kisumu, Kenya; Safe Water and AIDS Project, Kisumu, Kenya

## Abstract

**Background:** Various milestones have been set targeting the elimination of schistosomiasis with the vision of “a world free of schistosomiasis” including the 2020 and 2025 goals. Despite the increased control and prevention efforts, schistosomiasis still affects many people, partly because treatment doesn’t cover all at-risk populations. Preventive chemotherapy (PC), the cornerstone of control interventions is primarily inclined toward school-age children (SAC), whose prevalence also informs interventions for other age groups in the community. Whereas prevalence in SAC has been shown to be a good indicator of the prevalence in other age groups, it remains to be seen as if this is not true in different epidemiological settings and risk areas.

**Methods:** This study evaluated the predictive value of schistosomiasis prevalence among SAC in estimating community-wide prevalence in Siaya County, Kenya. A single stool sample was collected from participants aged 2-50 years. Stool samples were collected from preschool-age children (2-6 years), SAC (7-14 years), adolescents (15 to <18 years), and adults (≥18-50 years) in a cross-sectional survey. The prevalence and intensity of *Schistosoma mansoni* infection were determined using the Kato-Katz technique (single stool, 4 slides) and compared across the age groups and risk categories (low >0 to <10%; moderate ≥10% to <50%; and high (≥50%).

**Results:** Of the 1,814 participants sampled, 25.6% (95% CI: 23.7-27.7) were infected with *S. mansoni*. There was no significant difference between the mean egg intensity of *S. mansoni* in SAC compared to the other age groups. There was a strong positive correlation between SAC prevalence and intensity and community-wide prevalence and intensity; r_s_ =0.8, P<0.001, and r_s_ =0.6, P<0.001 respectively. A positive relationship between the overall *S. mansoni* prevalence and *S. mansoni* prevalence in SAC was observed. The regression analysis indicated that SAC prevalence/intensity can be used to predict overall *S. mansoni* prevalence/intensity. In terms of age groups, it can predict in all age groups but adolescents.

**Conclusions:** In this setting, generally, the prevalence of *S. mansoni* among SAC was a good predictor of the prevalence in other age groups and the overall (all age groups combined) prevalence. However, in terms of risk strata SAC data was only a good predictor of overall prevalence and intensity only in the moderate risk stratum. These findings suggest that *S. mansoni* prevalence and intensity among SAC are valid for community sampling purposes and informing interventions including MDA at the community level.

## Introduction

Schistosomiasis (Bilharzia) is caused by parasitic flatworms of the genus *Schistosoma* and affects approximately 220 million people worldwide with 90% of the burden concentrated in sub-Saharan Africa (SSA) (1). According to the World Health Organization (WHO), approximately 652 million people are at risk of the infection with about 200,000 deaths being reported annually. This infection exerts a great health and financial burden on the economies affected (2). In Kenya the disease is endemic in 62 out of 290 sub-counties and infects approximately 9 million people and about 17 million are at risk (3). The 3 main endemic regions are the coastal region *(Schistosoma haematobium*), parts of central and lower eastern areas (both *Schistosoma mansoni* and *Schistosoma haematobium),* and Lake Victoria basin area (mainly *Schistosoma mansoni*).

Success in schistosomiasis control is widely recorded in many countries with significant decrease in both prevalence and infection levels (4). However, in SSA this enervating infection remains a burden due to various factors that are responsible for continuous and persistent transmission. These include amongst others climatic changes, occupational activities, and proximity to water bodies (2). This infection is more prevalent in poor rural settings where fishing and agriculture are the main occupations (5). Poor water, sanitation and hygiene (WASH) is one of the key factors affecting the transmission of schistosomiasis. Infections occur during routine agricultural domestic occupational (like washing, bathing and fetching water) and recreational activities such as swimming, etc. These activities expose individuals to waters infested with the intermediate host (snail). School-age children (SAC), adolescents, women and young adults carry the greatest burden of morbidity and mortality associated with this infection(6–8). Equally, young children under 5 years are exposed especially in endemic regions. This mainly occurs when caregivers carry them along to the waterbodies (9, 10).

The WHO 2011 preventive chemotherapy (PC) strategy recommends that data from SAC can be used to assess community prevalence and that mass drug administration (MDA) should be conducted based on this information (11, 12). To date, monitoring and treatment of schistosomiasis is based on data from SAC only and very little attention is given to these other groups who are equally at risk. Integrating other age groups into the control programs will help accelerate the progress of control and elimination of schistosomiasis (13). The global community discusses strategies for incorporating other age groups especially adults and pre-school age children (PSAC) into treatment and monitoring plans. However, one of the main questions that remain unanswered is whether the WHO recommended prevalence among SAC is a good predictor of prevalence in other age groups in different epidemiological settings with varying risks of transmission. Control strategies for schistosomiasis have largely relied on prevalence data from SAC, which has also selectively excluded other at-risk groups such as PSAC and adults from interventions. This systematic exclusion of other at-risk groups contributes to the thriving of reservoirs of infection which will hamper the achievement of the WHO 2030 global elimination targets for schistosomiasis (14, 15).

Although WHO recommends that prevalence in SAC can be used to infer prevalence in other age groups (11, 16, 17), this was not the case for one other setting (18). This study aimed at evaluating a strategy that can be used to quantify the prevalence and intensity of schistosomiasis for other age groups and the community as a whole using a predictive index from SAC data and also in different risk regions (low, moderate, and high).

## Materials and methods

### Research population and design

The cross-sectional study was carried out in Siaya County, bordering Lake Victoria in western Kenya. The region is endemic for schistosomiasis, with an inverse relationship between the prevalence of infection and distance from the lake (19) Many rounds of mass drug administration (MDA) with praziquantel have been conducted especially on SAC (20, 21). Considering that the main economic activities in the area are fishing and agriculture, other age groups such as adolescents and adults and adolescents are exposed to infection. In addition, young children (under 5 years) are infected when their caregivers carry them along to the water bodies during domestic chores such as laundry.

### Sampling procedure and sample size determination

Stratified sampling was used to classify the eligible population into 4 strata depending on age group (PSAC 2-6 years, SAC 7-14 years, adolescent 15-18 years & adults >18 years). Study villages were divided into 3 risk strata according to the WHO-defined risk thresholds for schistosomiasis (low risk >0 to <10%, moderate risk ≥10% to <50%, and high risk ≥50%). The survey was conducted in 29 purposively selected villages. The total estimated population (N) per age group in these selected villages is 870 (an average of 30 per age group per village). A design effect (DEFF) of 1.58 (22) was applied.

Slovin’s 1960 formula (23) was used to calculate the sample size for each age group per village with an expected refusal rate of 15%. This gave n = 498 participants per age group (giving approximately 17 per age group per village). Before data collection, consent/assent was obtained for each participant. Parental/guardian consent was also obtained for each child. Participants were excluded from the study if they didn’t provide assent or consent or were observably ill or had an underlying medical condition, as judged by the study nurse.

### Data collection instruments

Data was collected using specially designed forms with the Commcare application (Dimagi Inc.) installed on Android smartphones which was then uploaded to a dedicated Commcare server. Laboratory data were recorded in a lab booklet/questionnaire and later entered via smartphones (Commcare application) into the same database.

### Reliability, validity and quality control

To ensure reliability, stool sample evaluation was performed by trained microscopists who were blinded to the villages where the samples were collected. To warrant the validity of the instruments, the questionnaire (developed in the Commcare application) was designed taking into account validation and logical aspects, especially for questions involving skip patterns. In addition, the questions were administered in a simple way for ease of understanding by participants. To improve diagnostic sensitivity for the Kato-Katz technique, 4 slides were prepared from each stool sample. Eggs in stool samples were counted by two independent microscopists and any discrepancy in the results of the two was reconciled by comparing them to the results of a third independent microscopist.

### Stool sample collection and processing

A single stool sample was collected from each participant. The stool samples were tested using the Kato-Katz technique (duplicate 41.7 mg fecal) for detection of *S. mansoni.* Four slides were prepared from the single stool sample and the slides were allowed to clear for about 24 hours before microscopic examination. The laboratory technicians counted the number of eggs and recorded them in the lab booklet. This data was then entered into the Commcare application using smartphones. At the analysis level, the intensity of *S. mansoni* was enumerated and expressed as eggs per gram (epg) of feces. Positive participants were grouped into intensity levels, low(1-99epg), moderate (100-399epg), and heavy(≥400epg).

### Data analysis

Data cleaning and analysis were done using STATA version 14.0 (STATA Corp. LP., College Station, TX, USA). The normality test was conducted using the Shapiro-Wilk test and histogram plots. The arithmetic means egg counts calculated from the 4 slides (24)data were multiplied by 24 to convert the egg counts to eggs per gram (epg). For each participant, stool results were used to determine the presence of *S. mansoni* and its intensity. For each village, the *S. mansoni* prevalence, and arithmetic mean (AM) epg were calculated, as well as the prevalence for each age group. Villages were classified into risk strata according to the prevalence of *S. mansoni* using the WHO recommended criteria (low risk >0 to <10%, moderate risk ≥10% to <50%, and high risk ≥50%) and analysis to come up with a regression equation conducted for each stratum. Kruskal Wallis test was used to compare the prevalence of the 4 age categories and a post hoc test (Dunnett’s test) was conducted to compare prevalence in SAC with each of the other 3 categories. Spearman’s correlation (r_s_) analysis was conducted, and scatter plots were plotted to illustrate the relationship between overall prevalence and intensity as a function of SAC prevalence and intensity. Linear regression analysis was conducted to come up with a predictive model Y = a+bX (where Y is overall prevalence/intensity or prevalence/intensity of other age groups and X = prevalence/intensity of school-age children). All statistical tests and confidence intervals were conducted at a 5% level of significance.

### Ethical approvals

Ethical approval for this study was obtained from the Board of Postgraduate Studies of Jaramogi Oginga Odinga University of Science and Technology (H153/4268/2019). This study was nested within a larger study whose ethical approval was obtained from the Maseno University Ethics Review Committee (MSU/DRPI/MUERC/00675/19). Additional approval for the larger study was obtained from the Kenya National Commission for Science, Technology and Innovation (NACOSTI/P/75695/30230). Parents and participants were provided with information sheets detailing the purpose of the study and what to expect. Written informed consent by parents or guardians and verbal assent by participating children were obtained before enrolment in the study.

## Results

A total of 29 villages were involved in the study with 1,814 participants enrolled. An almost even proportion by gender was represented, 45% (818) for males and 55% (996) for females. The mean age was 17.9 years. The enrollment was 23.7% (430) for PSAC, 27.7% (502) for SAC, 21.6% (392) for adolescents, and 27.0% (490) for adults.

The helminth egg count data was tested for normality before analysis and non-parametric tests were subsequently employed for analysis.

### Overall prevalence and intensity of helminth infections

Overall, 30.5% (548) of the participants were infected either with *S. mansoni* or one of the soil-transmitted helminths (STH), while 18.0% (324) of the participants were infected with one or more STH species. The most prevalent helminth infection among the participants was *S. mansoni* 25.6% (465) as shown in (**Table 1**).

**Table 1.** Overall (all age groups combined) prevalence and intensity of infection for *Schistosoma mansoni* and soil-transmitted helminths (STH) in Siaya county, western Kenya.

| Species<br>Infection | Overall<br>Prevalence %, (Range) | Intensity Threshold Prevalence |  |  | Intensity(arithmetic mean epg ± SD) |
| --- | --- | --- | --- | --- | --- |
|  |  | Light | Moderate | Heavy |  |
| Any helminth | 30.5 (28.4-32.6) | 21.3 | 5.5 | 3.3 |  |
| *S. mansoni* | 25.6 (23.7-27.7) | 16.9 | 5.4 | 3.3 | 215.4±628.8 |
| Any STH | 18.0(16.3-19.9) | 17.9 | 0.1 | 0.0 |  |
| Hookworms | 0.1(0.02-0.4) | 0.1 | 0.0 | 0.0 | 102.0±84.9 |
| *Ascaris lumbricoides* | 0.2(0.00-0.1) | 0.1 | 0.1 | 0.0 | 4536.0±7742.5 |
| *Trichuris trichiura* | 17.9(16.2-19.7) | 17.8 | 0.1 | 0.0 | 24.7±207.5 |

The proportions of individuals infected with multiple helminth species were slightly higher than those infected with *S. mansoni* only.

### Prevalence and intensity of *Schistosoma mansoni* by age group

We assessed the overall prevalence and the prevalence for each *S. mansoni* infection intensity level segregated by age group. Most of the *S. mansoni* infections were of light intensity (100-399 epg, with heavy-intensity infections (≥400 epg) accounting for the least proportion (**Table 2**). The prevalence for light and moderate infection levels was highest among adults followed by adolescents. However, the mean eggs per gram were highest among Preschool-age children 353.0(88.7-617.2).

**Table 2.** Prevalence and intensity levels of *Schistosoma mansoni* per age group.

|  | Positive | Light | Moderate | Heavy |  |
| --- | --- | --- | --- | --- | --- |
| Age group | n (%) | (1-99 epg) | (100-399 epg) | (≥400 epg) | Mean epg* (95% CI) |
| PSAC | 83 (19.3) | 12.8 | 3.0 | 3.3 | 353.0 (88.7-617.2) |
| SAC | 112 (22.3) | 14.1 | 6.0 | 2.2 | 215.3 (113.0-317.6) |
| Adolescent | 114 (29.1) | 19.6 | 5.4 | 4.1 | 173.1 (116.5-229.7) |
| Adults | 156 (31.8) | 21.0 | 6.9 | 3.9 | 172.5 (116.1-228.8) |

### Predictive value of *Schistosoma mansoni* prevalence and egg intensity in school-age children to that of PSAC, adolescents, and adults in low, moderate, and high-risk communities

Villages were categorized into 3 risk strata (low-risk >0 to <10%, moderate-risk ≥10% to <50%, and high-risk ≥50%). There were 5, 20, and 4 villages in the low, moderate, and high risk strata, respectively.

As expected, the prevalence of *S. mansoni* was highest in the high-risk areas, with an overall prevalence of 63.2%. Results of Dunnett’s test revealed no statistically significant difference between the mean egg intensity (epg) of SAC compared with that of PSAC, adolescents or adults, all with P > 0.05 (**Table 3**).

**Table 3.** Pairwise comparison of mean epg for SACs versus other age groups.

| Age group | Contrast | Std. Err. | t | P value | 95% CI |
| --- | --- | --- | --- | --- | --- |
| SAC vs PSAC | 19.2 | 21.8 | 1.0 | 0.704 | (-31.8-70.4) |
| SAC vs Adolescent | 2.3 | 22.3 | 0.1 | 0.999 | (-50.1-54.7) |
| SAC vs Adults | 6.9 | 21.0 | 0.3 | 0.977 | (-42.5-56.3) |

### Correlation analysis

There was a positive relationship between the overall *S. mansoni* prevalence and *S. mansoni* prevalence in school-age children (r_s_ = 0.8, P <0.001) (**S1 Fig**). There was also a positive linear relationship between community-wide (all groups combined) mean egg intensity and SAC mean egg intensity (r_s_ = 0.6, P <0.001)

Spearman’s rank correlation analysis was conducted to assess the association between village-level prevalence and mean egg intensity for SAC and other age groups. There was a positive correlation between *S. mansoni* prevalence in SAC and that of other individual age groups as follows; PSAC (r_s_ = 0.5, P<0.001), adolescents (r_s_ = 0.6, P < 0.001), and marginally significant for adults (r_s_ = 0.4, P = 0.06). In terms of risk strata, SAC prevalence had a positive correlation with overall prevalence (all groups combined) only in the moderate risk strata (r_s_ = 0.7, P < 0.001). There was a positive relationship between *S. mansoni* mean egg intensity in SAC and overall mean egg intensities; PSAC (r_s_ = 0.4, P = 0.03) and adults (r_s_ = 0.5, P = 0.01) but not adolescents. A similar pattern was observed in the overall prevalence in the moderate and high prevalence strata; PSAC (r_s_ = 0.5, P = 0.04) and adults (r_s_ = 1.0, P < 0.001).

There was a strong positive correlation between SAC prevalence and intensity and community-wide prevalence and intensity; r_s_ = 0.8, P < 0.001 and r_s_ =0.6, P < 0.001 for overall prevalence and intensity respectively.

There was a positive linear relationship between the prevalence of *S. mansoni* in SAC and that of other individual age groups. As illustrated in (**S2 Fig**).

### Regression Analysis

Linear regression analysis was used to come up with a model that can predict the prevalence and intensity of *S. mansoni* in SAC to the other age groups (PSAC, adolescents, and adults) and the overall community. This analysis was conducted overall (all risk strata combined) and per risk strata (low, moderate and high). A regression model of **Y = a+ bX** where X is the prevalence of school-age children and Y is either the prevalence of PSAC, adolescents, adults, or overall community prevalence.

**Table 4:**
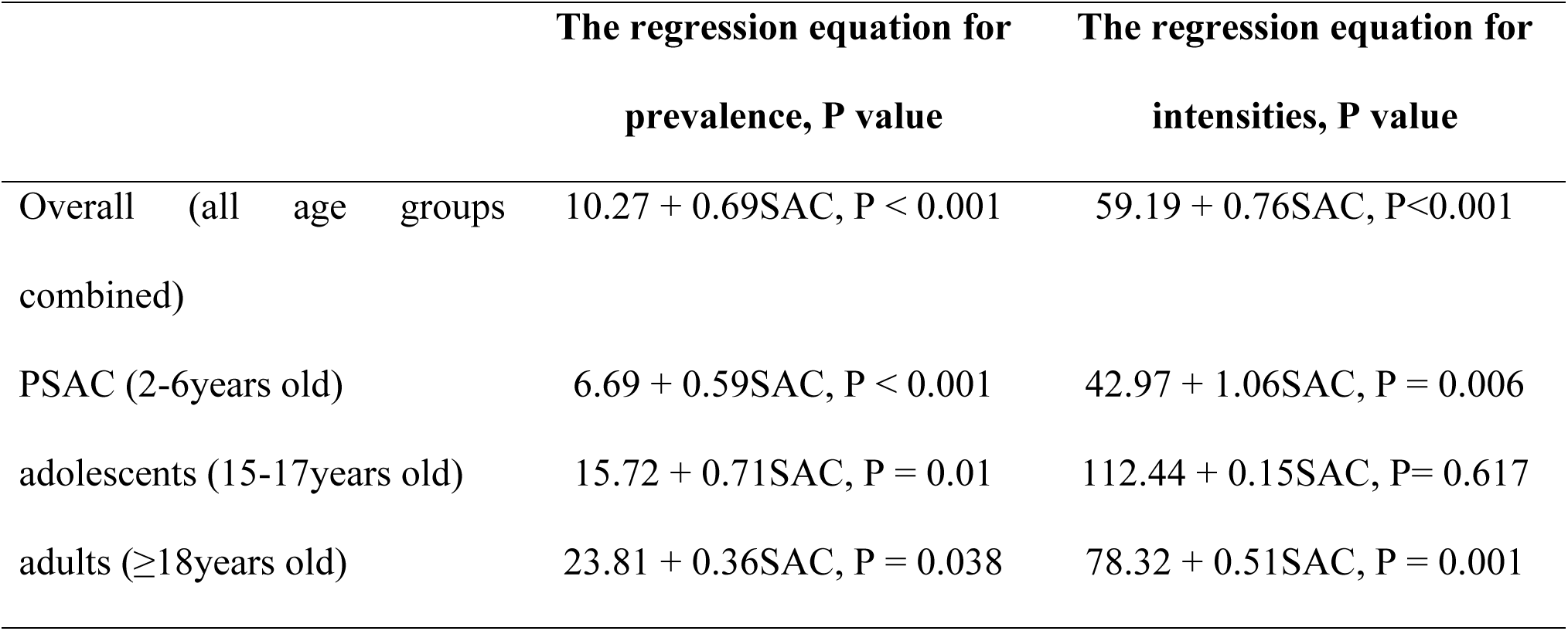
Regression equations for overall prevalence and mean egg intensities.

|  | The regression equation for prevalence, P value | The regression equation for intensities, P value |
| --- | --- | --- |
| Overall (all age groups combined) | $10.27 + 0.69SAC, P < 0.001$ | $59.19 + 0.76SAC, P < 0.001$ |
| PSAC (2-6years old) | $6.69 + 0.59SAC, P < 0.001$ | $42.97 + 1.06SAC, P = 0.006$ |
| adolescents (15-17years old) | $15.72 + 0.71SAC, P = 0.01$ | $112.44 + 0.15SAC, P = 0.617$ |
| adults ($\geq 18$years old) | $23.81 + 0.36SAC, P = 0.038$ | $78.32 + 0.51SAC, P = 0.001$ |

Results from linear regression modeling are as in **Table 5** below.

**Table 5:** Prevalence of *S. mansoni* infection in school-age children and their corresponding values in other age groups according to the regression equations.

| SAC prevalence (%) | Overall community-wide prevalence (%) | PSAC (%) | Adolescents (%) | Adults (%) |
| --- | --- | --- | --- | --- |
| 10 | 17.17 | 12.59 | 22.82 | 27.41 |
| 20 | 24.07 | 18.49 | 29.92 | 31.01 |
| 30 | 30.97 | 24.39 | 37.07 | 34.61 |
| 40 | 37.87 | 30.29 | 44.12 | 38.21 |
| 50 | 44.77 | 36.19 | 51.22 | 41.81 |
| 60 | 51.67 | 42.09 | 58.32 | 45.41 |
| 70 | 58.57 | 47.99 | 65.42 | 49.01 |
| 80 | 65.47 | 53.89 | 72.52 | 52.61 |
| 90 | 72.37 | 59.79 | 79.62 | 56.21 |
| 100 | 79.27 | 65.69 | 86.72 | 59.81 |

The above findings were all statistically significant, indicating that the prevalence of *S. mansoni* in school-age children can be used to predict the prevalence of *S. mansoni* in PSAC, adolescents, adults, or the overall community-wide. SAC mean egg intensity was also a predictor for mean egg intensity in PSAC, adults, and the entire community but not for adolescents.

In terms of risk strata, school-age data was a good predictor for overall prevalence and intensity in the moderate-risk regions (P = 0.012 and P < 0.001 respectively). It was also able to significantly predict the intensity of *S. mansoni* among adolescents in low-risk regions (P = 0.001).

**Table 6:** Regression equations for overall prevalence and mean egg intensities per risk strata.

|  | The regression equation for<br>prevalence, P value | The regression equation for<br>intensities, P value |
| --- | --- | --- |
| **Low-risk stratum** |  |  |
| Overall (all age groups<br>combined) | 7.64 - 0.347SAC, P = 0.391 | 34.03+ 0.79SAC, P = 0.403 |
| PSAC (2-6years old) | 15.02 - 1.04SAC, P = 0.494 | 60.01 - 1.14SAC, P = 0.395 |
| adolescents (15-17years old) | 1.21 - 0.06SAC, P = 0.987 | -15.24 + 3.01SAC, P = 0.001* |
| adults ($\geq 18$ years old) | 16.02 - 1.50SAC, P = 0.092 | 22.48 + 0.55SAC, P = 0.528 |
| **Moderate-risk stratum** |  |  |
| Overall (all age groups combined) | 16.91 - 0.379SAC, P = 0.012* | 76.35 + 0.49SAC, P < 0.001* |
| PSAC (2-6 years old) | 10.90 + 0.27SAC, P = 0.180 | 88.23 + 0.38SAC, P = 0.304 |
| adolescents (15-17 years old) | 26.90 + 0.31SAC, P = 0.468 | 149.79 - 0.24SAC, P = 0.554 |
| adults ($\geq 18$ years old) | 37.47 - 0.25SAC, P = 0.449 | 92.48 + 0.12SAC, P = 0.079 |
| **High-risk stratum** |  |  |
| Overall (all age groups combined) | 34.76 - 0.40SAC, P = 0.325 | -0.22 + 1.33SAC, P = 0.069 |
| PSAC (2-6 years old) | 56.17 - 0.004SAC, P = 0.998 | -389.18 + 3.26SAC, P = 0.192 |
| adolescents (15-17 years old) | 7.64 + 0.85SAC, P = 0.104 | 92.10 - 0.67SAC, P = 0.311 |
| adults ($\geq 18$ years old) | 60.51 - 0.05SAC, P = 0.815 | 227.97 + 0.52SAC, P = 0.77 |

## Discussion

This study was key in answering the question of whether SAC is a suitable reference group for predicting community-wide prevalence and more so in different risk areas. Such a predictor should be representative of the MDA target group and the entire community. School children are easily accessible from schools and this eases the process of sample collection. Both logistic and economic aspects need to be considered. Proper planning of control and intervention strategies for schistosomiasis requires information on community-level or local prevalence and mapping of the distribution of the disease. This information is critical for control programs. Whereas it is impractical and illogical to conduct assessment surveys on the infection in all possibly endemic communities, model-based statistics can help predict the prevalence and determine appropriate intervention master plans (25).

This study was conducted in Siaya County, a region that borders Lake Victoria. Previous studies have shown that these areas that border Lake Victoria are highly endemic for schistosomiasis (10, 26). In agreement with these studies, this study reported a significantly high prevalence of *S. mansoni* infection. However, the prevalence was marginally relatively higher among adolescents and adults as compared to other age groups, a pattern that was observed in other settings (27) (6). The slightly higher prevalence among adolescents and adults relative to SAC in the current study setting may be explained by differences in their daily activities and occupations that expose them to frequent water contact such as fishing, washing among others. For adults in particular, it may also be a result of the cumulative effects of years of exclusion from MDA conducted in Kenya by the National School-Based Deworming Programme (NSBDP) which only targets school-age children (28).

A positive linear relationship was observed between the prevalence and mean epg for SAC (7-15 years) and that of the entire community (all age groups combined). Moreover, there was no significant difference in the mean epg for SAC and that of other individual age groups, and a strong positive correlation between the prevalence of *S. mansoni* among SAC and that of the overall community. The linear regression model also confirmed that indeed SAC is a good predictor for overall prevalence and mean intensities for each age group and the entire community. These findings add to the evidence that SAC is indeed a suitable reference group and also confirm the WHO recommendation of using school-age children data to predict community-wide prevalence (29–31). In a study conducted in western Kenya, SAC (9-12 years) was a good predictor for community-wide prevalence and intensity of *S. mansoni* (11). Prevalence of *S. haematobium* among SAC (7-14 years) sufficiently estimated the community-wide prevalence in Mali (16). Similarly, a study conducted among 180,000 people from 3 municipalities in Brazil, found that the prevalence of schistosomiasis among children aged 7-14 years had a strong relationship with entire community prevalence (32). Another nationwide study by Guyatt revealed that SAC was a good and cost-effective predictive technique for community-wide prevalence that can be used to make intervention decisions not only at the district level but also at the national level (17). Additionally, another study conducted in an endemic area in Pernambuco reported that the prevalence of Schistosomiasis in SAC(6-15 years) is a suitable indicator for community prevalence(33).

There was a positive correlation between *S. mansoni* prevalence in SAC and that of other individual age groups but not significant for adults. In that regard this study’s regression analysis per risk strata indicated that *S. mansoni* prevalence in SAC was only a good predictor in moderate-risk areas with prevalence (≥10% to <50%) and not for low (>0 to <10%,) and high-risk areas (≥50%). That concluded that SAC data wasn’t a good predictor for adults and low and high-risk settings. Similar results were depicted in a study conducted in Nigeria (18) reporting that SAC was not an appropriate reference group for predicting prevalence among adults in high-risk regions where *S. mansoni* prevalence is ≥50%.

A limitation of this study is that the study was conducted in an area that has undergone several rounds of MDA targeting mainly SAC and no other age groups. It is therefore difficult to know if the findings can be inferred for an area where treatment has been provided across all age groups. Nevertheless, findings can be considered within the context and understanding that until recently, the recommended MDA strategy was premised on the fact that treatment is provided to a certain target group (in this case SAC), and that the benefits are accrued to all others age groups living in that endemic area. It is therefore reasonable to assume that any benefits accrued to SAC as a result of treatment are also experienced in the other age groups, as has been demonstrated for PSAC and adults(34). Additionally, existing evidence shows that SAC prevalence can overestimate prevalence in other age groups in the community, the degree of overestimation being dependent on the parasite species and the intensity level of infection (17, 35). This study considered only one parasite species (*S. mansoni*) which is the dominant species in the study area and did not include *S. haematobium.* Therefore, findings may only be readily applicable to *S. mansoni* and may suggest the need for careful considerations among other things, local transmission dynamics including parasite species.

## Conclusion

This study’s findings showed that the *S. mansoni* prevalence and intensity among SAC (7-15 years) was comparable to other age groups in the same epidemiological settings as well as with the entire community (all age groups combined). SAC is a good predictor/reference group for predicting community-wide prevalence and can be used to inform the planning and implementation of control interventions such as MDA. However, in terms of risk areas (categorized as low, moderate, and high risk), SAC was only a good predictor for schistosomiasis in moderate-risk communities.

Where resources are limited, predictive regression models may be of great value in generating the much-needed prevalence data to inform control strategies. Importantly, additional studies are required in different epidemiological settings with varying risks (low, moderate, and high), to ascertain the utility of SAC prevalence in informing community-wide prevalence.

## Data Availability

All raw data produced in the present study are available upon reasonable request to the authors

## Acknowledgments

This research was supported by the Safe Water and Aids project/KEMRI-CDC. We thank our colleagues who provided great insights and expertise that greatly made this research a success. We would also like to thank the study participants for voluntarily agreeing to take part in the research.

## Supporting Information

**S1 Fig.**
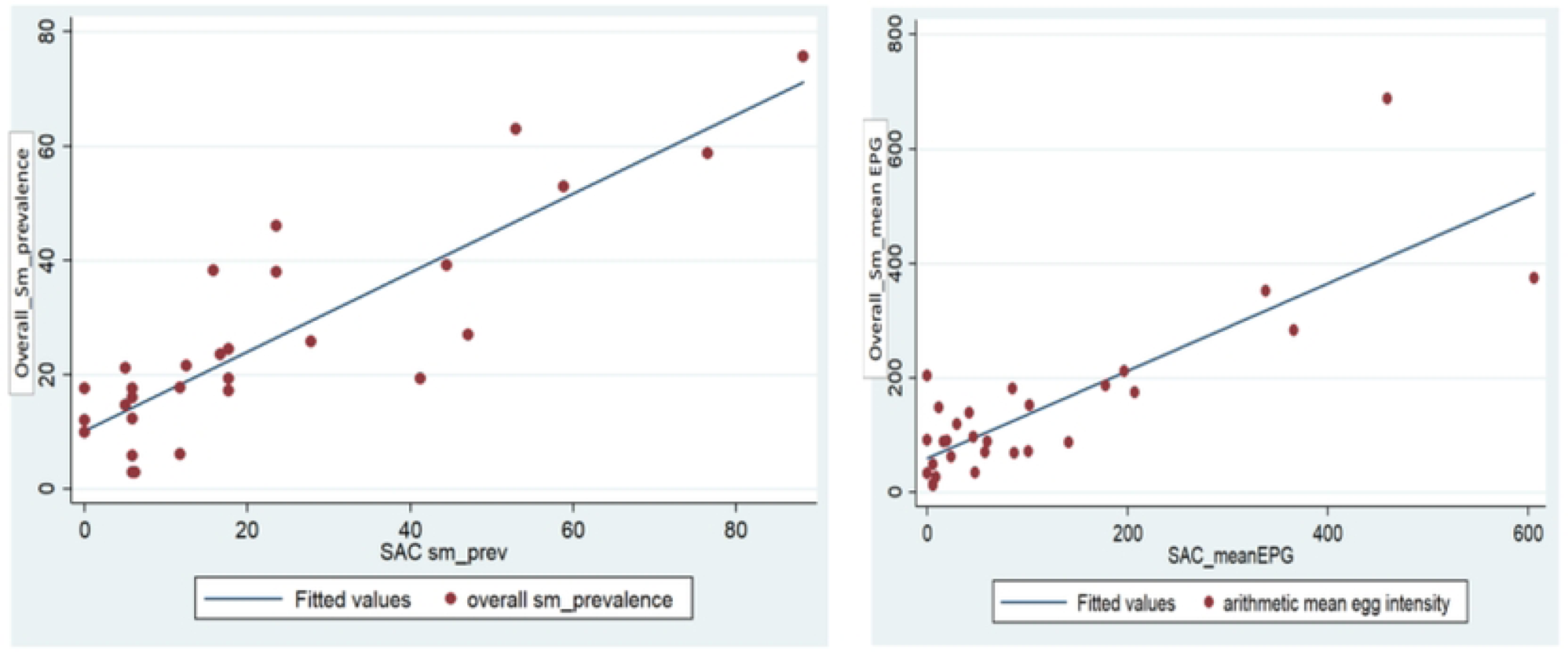
a. Scatter plot for overall prevalence and as a function of prevalence by village for school-age children b. Scatter plot for overall mean egg intensity as a function of mean egg intensity by village for school-age children

**S2 Fig.**
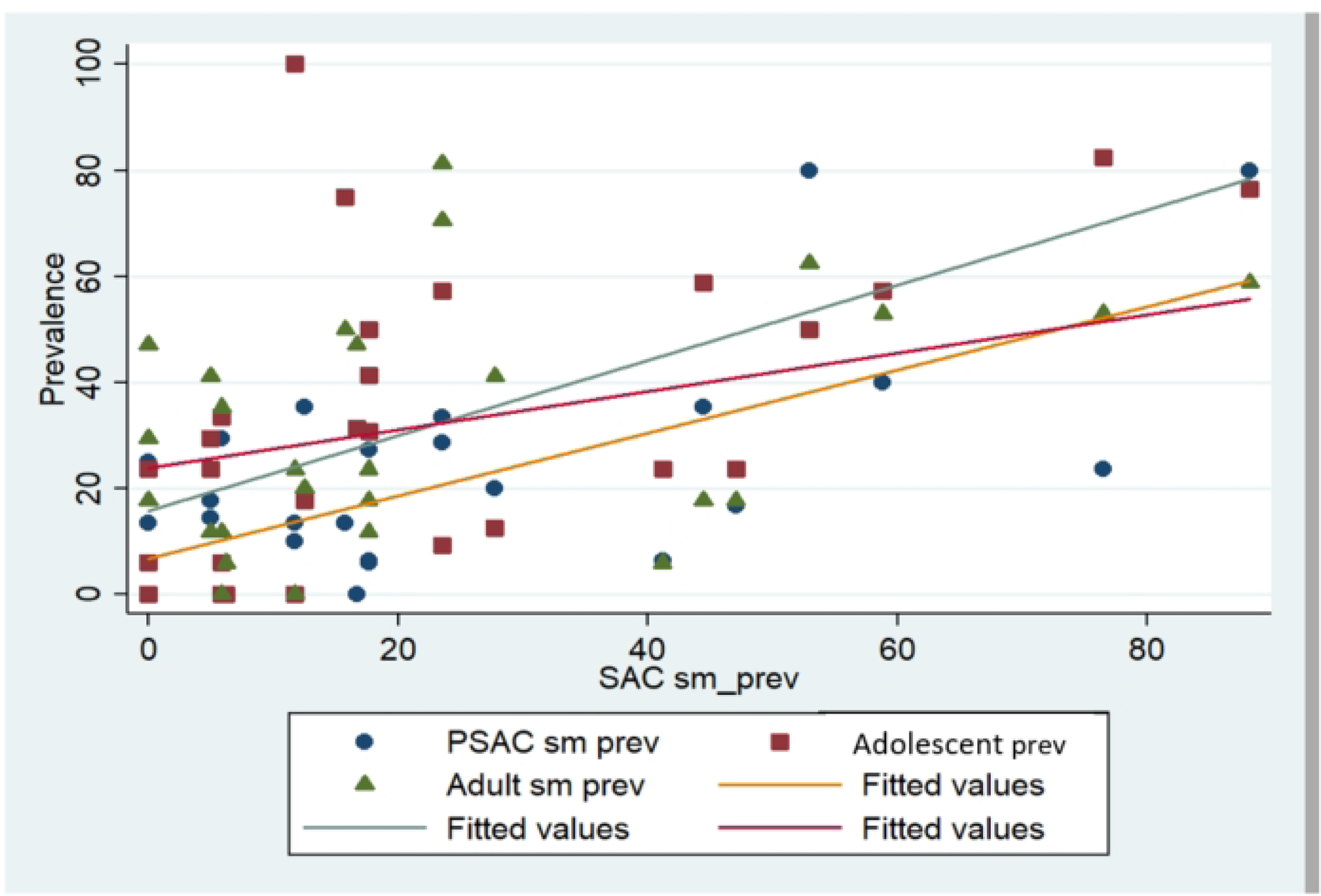
*Schistosoma mansoni* prevalence between age groups in Siaya County, western Kenya

